# Examining SARS-CoV-2 Interventions in Residential Colleges Using an Empirical Network

**DOI:** 10.1101/2021.03.09.21253198

**Authors:** Hali L. Hambridge, Rebecca Kahn, Jukka-Pekka Onnela

## Abstract

Universities have turned to SARS-CoV-2 models to examine campus reopening strategies^1–9^. While these studies have explored a variety of modeling techniques, all have relied on simulated data. Here, we use an empirical proximity network of college freshmen^10^, ascertained using smartphone Bluetooth, to simulate the spread of the virus. We investigate the role of testing, isolation, mask wearing, and social distancing in the presence of implementation challenges and imperfect compliance. Here we show that while frequent testing can drastically reduce spread if mask wearing and social distancing are not widely adopted, testing has limited impact if they are ubiquitous. Furthermore, even moderate levels of immunity can significantly reduce new infections, especially when combined with other interventions. Our findings suggest that while testing and isolation are powerful tools, they have limited benefit if other interventions are widely adopted. If universities can attain high levels of masking and social distancing, they may be able to relax testing frequency to once every two to four weeks.

## Introduction

When SARS-CoV-2 escalated to a pandemic in early 2020, universities and colleges around the world were forced to rapidly pivot to virtual instruction. Students were sent home and residential campuses were locked down as schools struggled to adapt to a new normal. As the pandemic continued into the summer, universities were faced with a difficult choice: reopen campuses with some return to traditional in-person instruction or continue teaching entirely online.

In the autumn of 2020, college administrators around the world turned to simulations to understand how enhanced public health protocols could mitigate the spread of SARS-CoV-2 on their campuses. Studies examined an assortment of preventive techniques and made different assumptions about compliance with their proposed policies ^1–9^. However, while researchers explored a variety of modeling techniques, from compartmental homogeneous mixing models to contact networks to agent-based models, all studies so far have only used simulated data. In their review of COVID-19 modeling studies in a university setting, Christensen et al. advocated for more research to be done using empirical mixing data ^3^. Here we take up that charge and examine how using a real-world contact network of students on a college campus, ascertained using smartphone Bluetooth data, changes our understanding of the role of repeat testing, isolation, and other strategies in the mitigation of SARS-CoV-2.

While several countries have recently begun to offer SARS-CoV-2 vaccines to their residents, the Council on Foreign Relations estimates that it will take years for the majority of the world’s population to become immunized against the virus ^11^. Furthermore, little is known about how long vaccine-induced immunity will last, whether it will protect against transmission, and how it will respond to more virulent strains of SARS-CoV-2, such as those that have emerged in early 2021. Yet, in spite of an unprecedented surge in COVID-19 cases in the United States and elsewhere, more colleges are making plans to reopen for the 2021 spring and fall semesters ^12^. While mitigation measures like testing and isolation can be costly and resource intensive, many colleges face dire financial straits if they fail to reopen their campuses.

## Methods

### Data

We analyze a close proximity interaction network from the Copenhagen Networks Study (CNS), which enrolled students from the Technical University of Denmark. Over 1, 000 students volunteered for the study, which examined multiple types of communication networks among this highly interconnected group ^13^. High-quality network data is rare, largely due to privacy concerns. As such, the researchers made privacy a central component of the study, providing participants with a platform to explore and visualize their data before consenting to its inclusion in the study ^10^. For this work, we focus exclusively on the Bluetooth proximity data from 706 students, which was made publicly available as of December 2019^10^.

Loaner smartphones were issued to study participants, who agreed to use the device as their primary phone for the duration of the study. Devices were configured to be Bluetooth discoverable at all times and to scan for nearby devices every five minutes. When scanning, a device emits a ping and receives responses from all nearby Bluetooth devices. Both devices then log this call and response, recording the device ID, timestamp, and an indicator of received signal strength (RSSI). This RSSI roughly correlates with physical distance; high values indicate close proximity while smaller values suggest the devices are distant or blocked by a physical barrier. Information on directionality was discarded so that the resulting data was symmetrical. Additionally, we removed empty scans and identifiers from devices not participating in the study. While the CNS followed students for several years, Bluetooth proximity data is only available for 28 days starting in February 2014.

The median number of proximity events per user over the course of the study was 10, 731 with a corresponding minimum of 1 and maximum of 30, 307. The mean was 11, 190.6 events per user, or about 400 events per day, with a standard deviation of 5, 131.3 or about 183 events per user per day. 5.9% and 11.2% of users had proximity event counts below 2800 (about 100 per day) and 5600 (about 200 per day) events, respectively, indicating perhaps not all participants were fully compliant in regularly using the study-issued device. The number of proximity events per user per day is shown in Figure 1.

**Figure 1:**
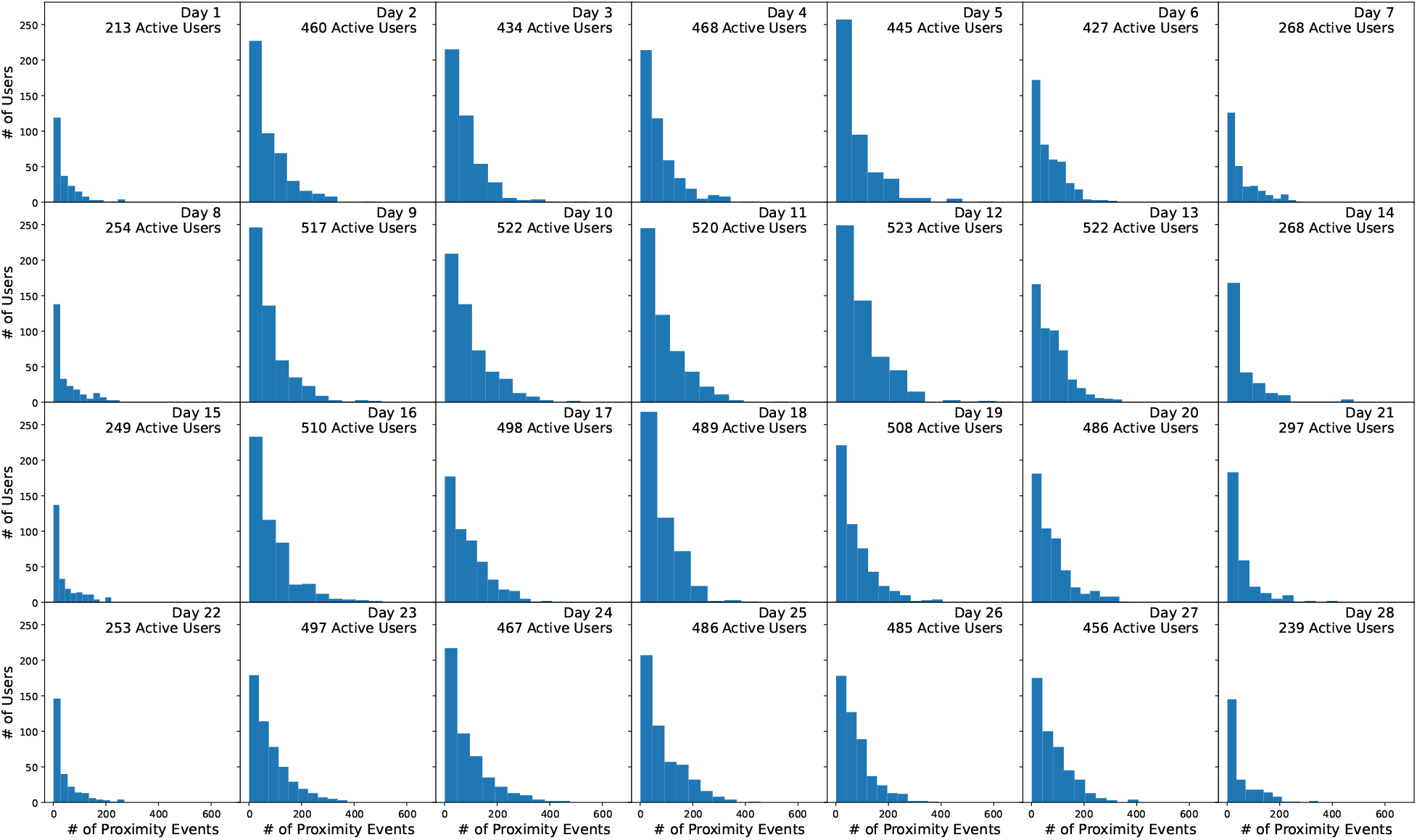
Number of proximity events per user per day for 675 students in the Copenhagen Network Study prox-imity data ^10^. Only close proximity Bluetooth connections (RSSI ≥ − 75, corresponding to physical proximity of approximately one meter) with participating devices are shown, corresponding to 8.0% of all detected proximity events. Number of active users indicates users with at least one Bluetooth ping with a fellow participants. All other users only had empty scans or pings with non-participating devices.

### Contact Network

Standard Bluetooth technology can detect devices as far as 5-10 meters away ^14^. However, since SARS-CoV-2 is generally only spread when individuals are near one another, we focused on close proximity interactions, excluding data with an RSSI value less than − 75 dBm. Previous studies have shown this RSSI threshold roughly corresponds to one meter ^14^.

We generated separate contact networks for each day of the simulation. For each day, we computed the number of close proximity interactions (i.e. RSSI ≥ − 75 dBm) between every pair of participants. Because devices were configured to scan every five minutes, a unique entry is recorded for every five minute time interval where a ping occurred. As such, the number of close proximity interactions between a pair of students is equivalent to the number of five minute intervals where a ping occurred. For instance, twelve interactions could represent one long continuous interaction that spanned an hour or several short intermittent interactions that took place over the course of a day. Both scenarios are interchangeable in our model, with the number of five minute intervals containing an interaction dictating the spread of the virus.

In order to model a typical university semester, we looped through the CNS data four times, thereby simulating a total of 16 weeks worth of interaction data. Thus, while the network structure varies from day to day, each four-week segment is identical. Although our approach does not capture seasonal trends in proximity, it does capture the stark contrast between weekdays and weekends. Also, the direction and strength of seasonal trends varies by latitude, which might need to be accounted for depending on the specific locale of the university one wishes to model.

### Epidemic Model

To model the spread of SARS-CoV-2 over the proximity network described in the previous section, we used a discrete-time stochastic susceptible-exposed-infectious-recovered (SEIR) compartmental model, shown in Figure 2. At each time step, individuals advance to the next compartment or remain in their current one probabilistically.

**Figure 2:**
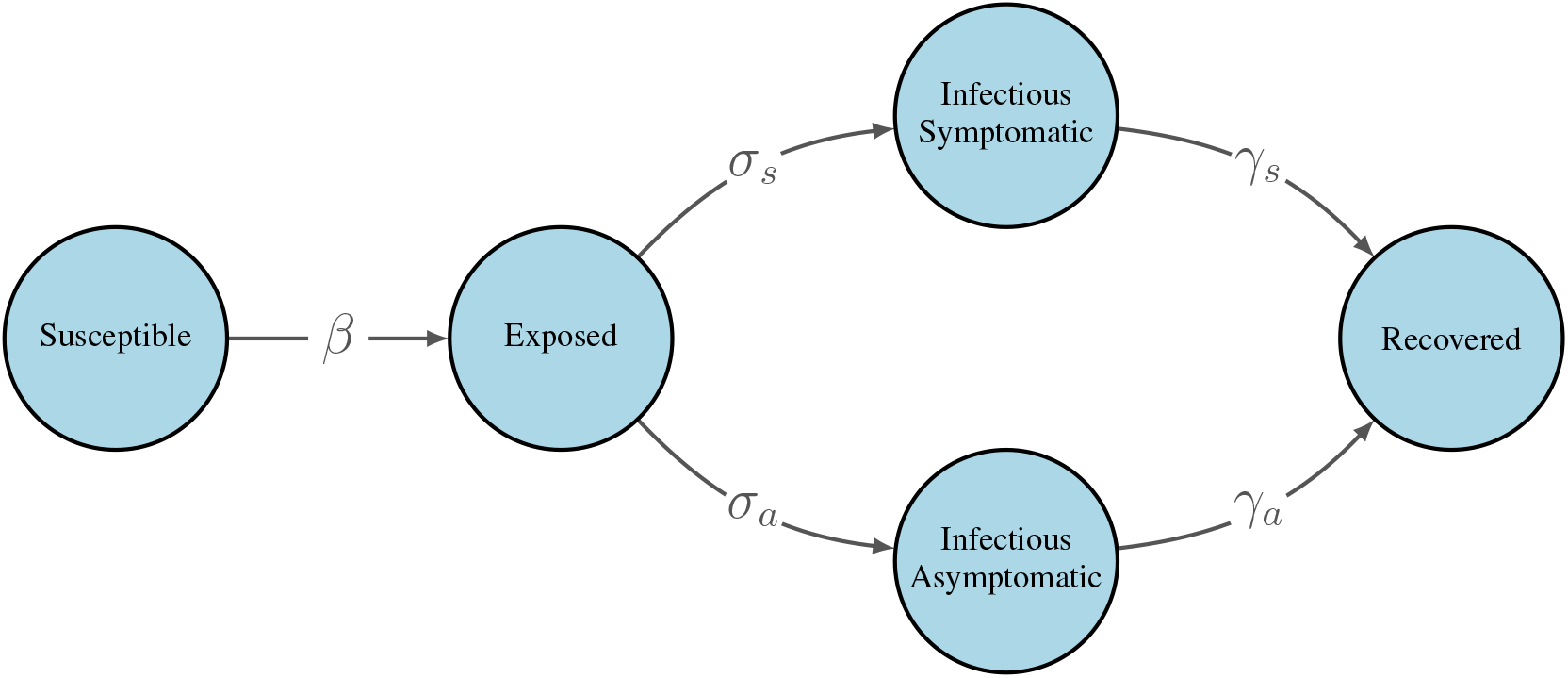
Modified SEIR model for the spread of SARS-CoV-2. *β* is a pair-specific transition probability per 5-minute close proximity interaction. All other transition probabilities are constant at each time step and do not incorporate the underlying contact network. For specific parameter values, see Table 1.

Because a significant fraction of COVID-19 patients are asymptomatic, estimated at 30.0% overall ^15^, we further divided the infectious compartment into symptomatic and asymptomatic subcompartments with separate transition probabilities into and out of these compartments. We allowed asymptomatic individuals to remain asymptomatic for the duration of their infection; that is, we did not assume all infected individuals eventually developed symptoms.

**Table 1:**
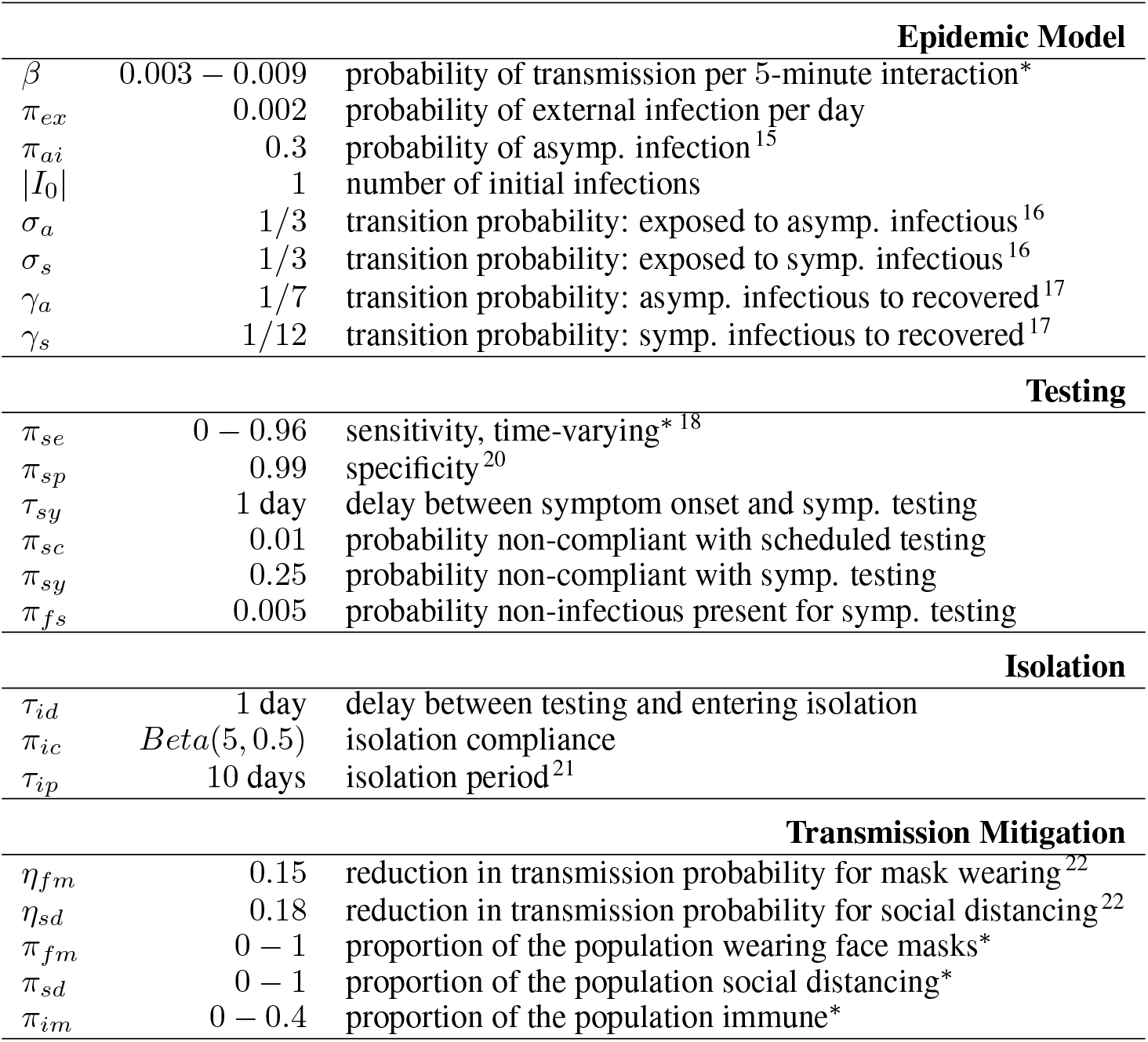
Parameters for simulation scenarios. Transmission probability, *β* is per 5-minute interaction with an infectious individual. All other epidemic transition parameters are per day, giving an average latent period of 3 days and an average infectious period of 7 and 12 days for asymptomatic and symptomatic infections, respectively. ^*\**^Transmission probabilities, proportion of the population wearing face masks, proportion of the population social distancing, and proportion of the population immune varied across simulations. Test sensitivity was time-varying. An overview of the epidemic model is shown in Figure 2. For additional details on parameter values, see the corresponding sections of the Methods.

Transition probabilities were structured as follows. At each time step, for each close proximity interaction, an infectious individual could transmit the virus to a susceptible individual with probability *β*. This parameter could be set to a universal value across all pairs of individuals, or be pair-specific, allowing for heterogeneous transmission probabilities that vary based on the pair’s precautions. Each interaction between an infectious individual and a susceptible individual was treated as an independent event, such that the probability of becoming exposed increased linearly with the number interactions with an infectious individual. Once an individual entered an exposed state, their symptom status was determined via Bernoulli trial. At each subsequent time step, exposed individuals transitioned to either a symptomatic infectious state or an asymptomatic infectious state with probabilities *σ*_*s*_ and *σ*_*a*_, respectively. We set *σ*_*s*_ = *σ*_*a*_ = *1*/*3*, leading to an average latent period of 3 days, consist with the literature ^16^. Infectious individuals then transitioned to a recovered state with probabilities *γ*_*s*_ and *γ*_*a*_ for symptomatic and asymptomatic individuals, respectively. Infectious times vary by disease severity, with mild to moderate cases infectious for no longer than 10 days after symptom onset and severe cases infectious for no longer than 20 days after symptom onset ^17^. As such, we selected *γ* values such that asymptomatic cases, which are inherently mild, had an average infectious period of 7 days while symptomatic cases had an average infectious period of 12 days. Note that in the case of symptomatic infection, this period includes a pre-symptomatic period where the student is able to infect others. If an individual did not transition to the next compartment at a given time step, they were considered for transition at each subsequent time step.

In addition to infection via the contact network, we also allowed each individual to develop infection due to outside exposure, i.e., an exposure acquired from the broader community, such as the urban environment within which the university is located. At each time step, all susceptible individuals had *π*_*ex*_ = 0.002 probability of becoming exposed, regardless of their contact network interactions.

### Testing

We considered symptomatic testing and scheduled testing in our simulations, and assumed both types of testing were done via polymerase chain reaction (PCR). Symptomatic testing occurs when a student seeks testing after experiencing symptoms. However, at each time step only a fraction of symptomatic individuals seek testing as inevitably a portion of the population will dismiss their symptoms. For those that do present for testing, we incorporated a short delay between symptom onset and testing. This delay could be attributed to difficulty in scheduling a testing appointment, the patient waiting to see if symptoms persist, or some combination of the two. Note that each infectious symptomatic individual only seeks testing once during their illness; if the individual tests negative, they do not pursue further symptomatic testing. In order to account for individuals who may experience non-COVID related flu-like symptoms, a fraction of non-infectious individuals also present for symptomatic testing at each time step. These uninfected individuals may present for symptomatic testing multiple times over the course of the semester.

Under scheduled testing, every member of the population is tested regularly in an effort to identify additional cases that would otherwise go undetected. At each time step, a fixed fraction of the population is tested. For instance, if the entire population is to be tested every 7 days, then 1*/*7 of the population is each tested each day of the week. Furthermore, individuals are always tested at regular intervals so that the time between tests is constant for each student. However, our model allows for a small fraction of the population to be non-compliant at each time step, thereby missing their scheduled test.

For both types of testing, test sensitivity was dependent on the time since exposure. To model this sensitivity, we used the nasopharyngeal swab data from Wikramaratna et al ^18^. However, because their model only included the time after symptom onset, we had to impute pre-symptomatic test sensitivity. Our model assumes that symptomatic infectious students develop symptoms two days after becoming infectious. Research has shown that viral load peaks at symptom onset, with similar loads pre- and post-symptom onset ^19^. As such, we mirrored the post-symptomatic sensitivity so that the two days prior to symptom onset had the same sensitivity as the two days after symptom onset. We assumed zero test sensitivity during the latent period and beyond the data available from Wikramaratna et al. We used this model for all students, regardless of symptom status, as studies have shown viral loads are comparable between symptomatic and asymptomatic patients ^19^. Figure 3 illustrates the functional form for our test sensitivity model. Specificity was fixed at 99% for all susceptible and recovered individuals, a widely reported specificity among COVID-19 tests approved for use by the U.S. Food and Drug Administration ^20^.

**Figure 3:**
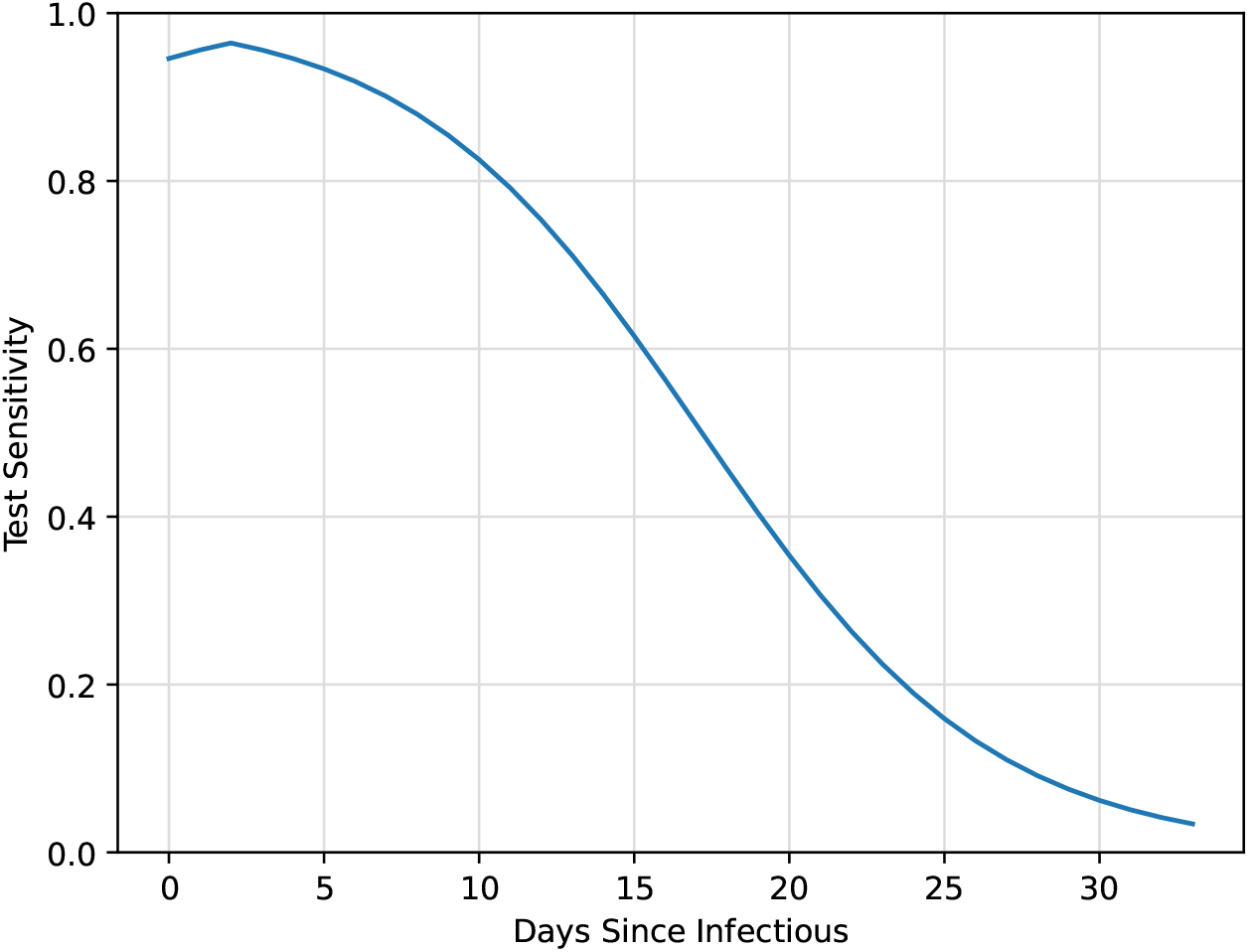
SARS-CoV-2 test sensitivity based on nasopharyngeal swab data adapted from Wikramaratna et al ^18^. Test sensitivity outside the range shown here was assumed to be zero.

### Isolation

Upon testing positive, individuals were considered for isolation, regardless of their true underlying disease state. Each individual was assigned a delay that dictated the number of days between testing positive and being eligible for isolation. This delay, which captures both the lag between testing and receiving results and the time needed to enter isolation, could be set to a universal value for all individuals or be individual-specific. Once an individual’s delay elapsed, they immediately entered isolation. However, individuals were also assigned an isolation compliance probability, drawn from a *Beta* (5, 0.5) distribution (mean: 0.91, standard deviation: 0.11). At each time step, each individual in isolation could either be deemed fully compliant or wholly non-compliant based on a Bernoulli trial using their compliance probability. If compliant, the individual remained in isolation, unable to infect others for that time step. If non-compliant, the individual would participate in their usual interactions during that time step and could infect others via their contact network. An independent trial was conducted each day the individual was in isolation. Isolation duration was set to 10 days based on the latest Centers for Disease Control and Prevention guidelines ^21^.

### Scenarios

Due to the stochastic nature of our model, each simulation was repeated 100 times to capture variability in the spread of the virus. This is particularly important given our underlying contact network; the location of initial infections can be highly influential in the ultimate course of the epidemic.

Overall parameter settings are summarized in Table 1. The time step for our simulations was set to one day; transition probabilities and other parameters were scaled to reflect this choice. For our first scenario, we set the transmission probability to a universal value for all individuals and explored low, medium, and high transmission scenarios, setting *β* = 0.003078, *β* = 0.006157, and *β* = 0.009235, respectively. The small magnitude of these transmission probabilities reflects the fact that they are per 5-minute interaction. These transmission probabilities roughly correspond to *R*_0_ values of 1.5, 3.0, and 4.5, though there is considerable variability in the frequency of contacts across both participants and days. For each of these transition probabilities, we examined scheduled testing frequencies of 3, 7, 14, and 28 days. While our model allowed for individual-specific values, the symptomatic testing delay and isolation delay were set to a single value for all individuals. Isolation compliance, however, varied across individuals as described previously.

### Transmission Mitigation

In addition to these low, medium, and high transmission scenarios, we also considered a setting where student behavior led to individual-specific transmission rates. Under this setting, each pair of student had a unique *β* value based on the interventions they had adopted. We specified a proportion of the population that would wear face masks, and randomly assigned a proportion of the population to abide by social distancing. Since homophily, the tendency for people to associate with others whom they perceive to be similar to themselves, is typically present in social networks ^23^, we hypothesized that friend groups might share similar views about COVID-19 and related mitigation efforts. We therefore considered both clustered and non-clustered assignment of mask wearing across the network, a scenario which cannot be studied with standard epidemiological models. Under non-clustered assignment, participants were randomly assigned to either follow one precaution, both precautions, or neither. To create clusters of mask users, we created a weighted contact network for the entire study period where each edge represented at least one contact between two participants. Each edge was weighted by the number of contacts over the entire course of the study. A set of 7 initial seed nodes were randomly chosen to wear face masks. We then “spread” mask wearing to their contacts where neighbors with more interactions had a higher probability of wearing a face covering. This process was iterated until the desired proportion of mask wearers was reached.

Based on the work by He et al. ^22^, we hypothesized that social distancing would reduce baseline transmission by a multiple of 0.18 and that each face covering would reduce baseline transmission by a multiple of 0.15. Thus, for each participant who wore a face mask, pair-wise transmission was reduced by 85%. Social distancing is symmetric in nature, so if one or both participants complied, their transmission was reduced by 82%. For this analysis, we used a baseline *β* = 0.006157.

We also performed a sensitivity analysis with reduced efficacy of masking and social distancing using alternative efficacies found in the literature. For each participant who wore a face mask, pair-wise transmission was reduced by only 68% ^24^. Similarly, social distancing reduced transmission by only 23% ^25^.

We considered the setting where a proportion of the population was immune, either due to previous illness or vaccination. Participants were randomly assigned to either be immune or susceptible. If a participant was set to be immune, they remained immune for the duration of the simulation and did not become ill or infectious.

Lastly, to examine the comparative effectiveness of the interventions we considered, we performed an ordinary least squares regression analysis for the moderate transmission scenario (*β* = 0.006157 or *R*_0_ ≈ 3.0). The dependent variable was cumulative incidence over the course of the simulated semester while the independent variables were testing frequency, proportion wearing face masks, proportion social distancing, and proportion immune. Because simulations were repeated multiple times, each combination of parameters had 100 realizations.

## Results

After removing non-participating devices and empty scans from the Bluetooth data, there were a total of 2, 426, 279 Bluetooth pings (44.3% of all pings) and 692 users (98.0% of all users), indicating 14 users did not have any proximity events with other study participants. The proximity networks for each of the 28 days of the study are shown in Figure 4. The networks have a large connected component on the weekdays when students are likely active on campus and attending classes. On the weekends, shown in the first and last columns of Figure 4, the networks are more loosely connected and fewer users are interacting with fellow study participants.

**Figure 4:**
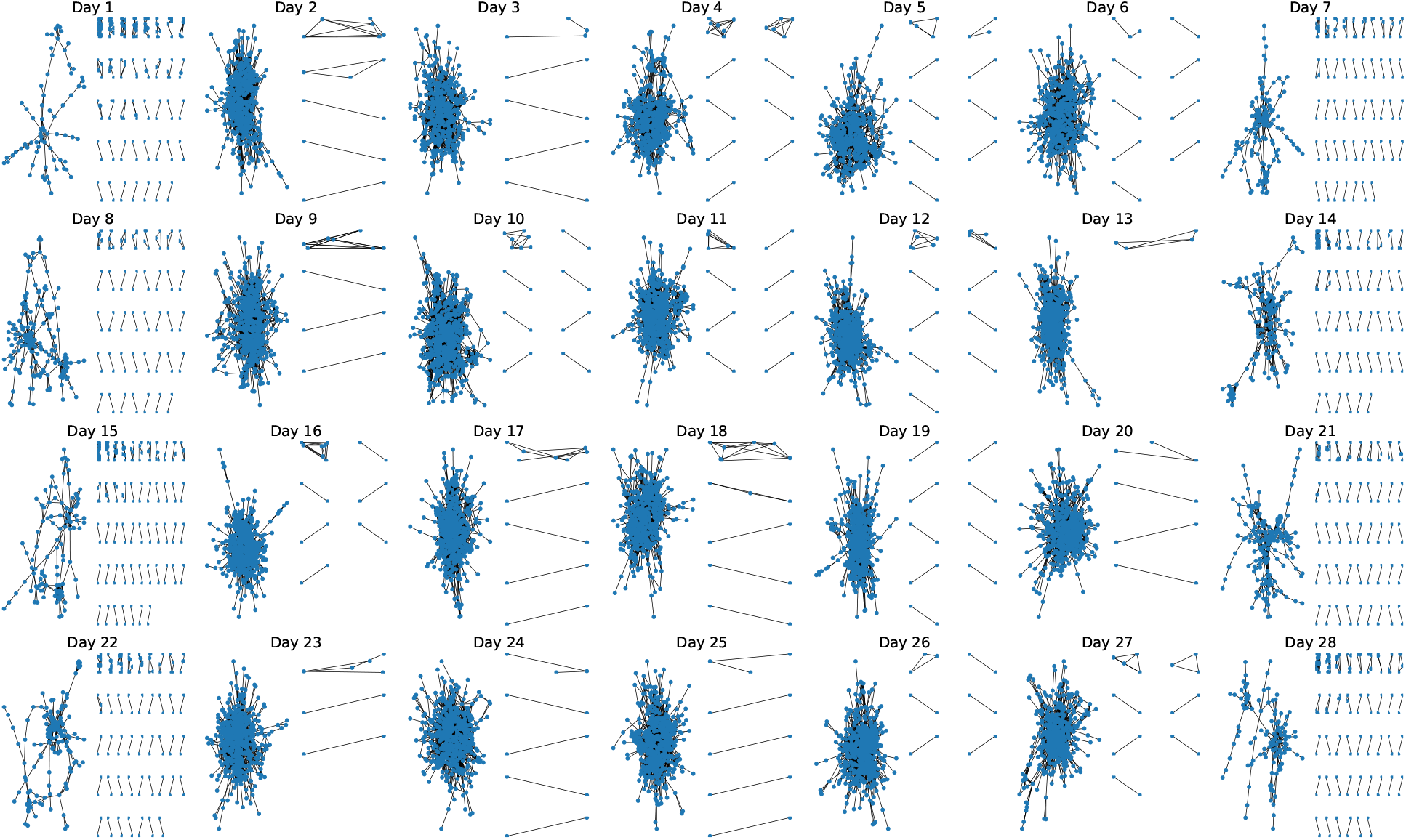
Proximity networks by day for 675 students. Each node represents a study participant and each edge represents the presence of one or more Bluetooth pings on that day. Only close proximity Bluetooth connections (RSSI ≥ − 75, corresponding to physical proximity of approximately one meter) with participating devices are shown. For each day, the largest connected component is shown at left, with the remaining connected components displayed at right ordered from largest to smallest.

Incidence for our low (*R*_0_ ≈ 1.5), moderate (*R*_0_ ≈ 3.0), and high (*R*_0_ ≈ 4.5) transmission scenarios are shown in Figure 5. Regardless of transmission levels, increased testing frequency reduced the number of infections observed over the course of the semester, though the effect was relatively small for the low transmission scenario. For *R*_0_ ≈ 1.5, testing every 3 days resulted in an average of 29.2% of students infected by the end of the semester, while testing every 7, 14, and 28 days gave rise to 31.8%, 36.4%, and 40.7% students infected, respectively (no testing: 53.2%). For *R*_0_ ≈ 3.0, testing every 3 days resulted in an average of 42.1% of students infected by the end of the semester, while testing every 7, 14, and 28 days gave rise to 47.4%, 54.9%, and 61.4% of students infected, respectively (no testing: 73.4%). Finally, for *R*_0_ ≈ 4.5, testing every 3 days resulted in 54.2% of students infected by the end of the semester, while testing every 7, 14, and 28 days gave rise to 60.3%, 67.3%, and 72.6% of students infected, respectively (no testing: 81.1%). Thus, while increased testing and subsequent isolation impacted the number of infections for a given transmission probability, ultimately reducing disease transmission had a greater impact on cumulative incidence.

**Figure 5:**
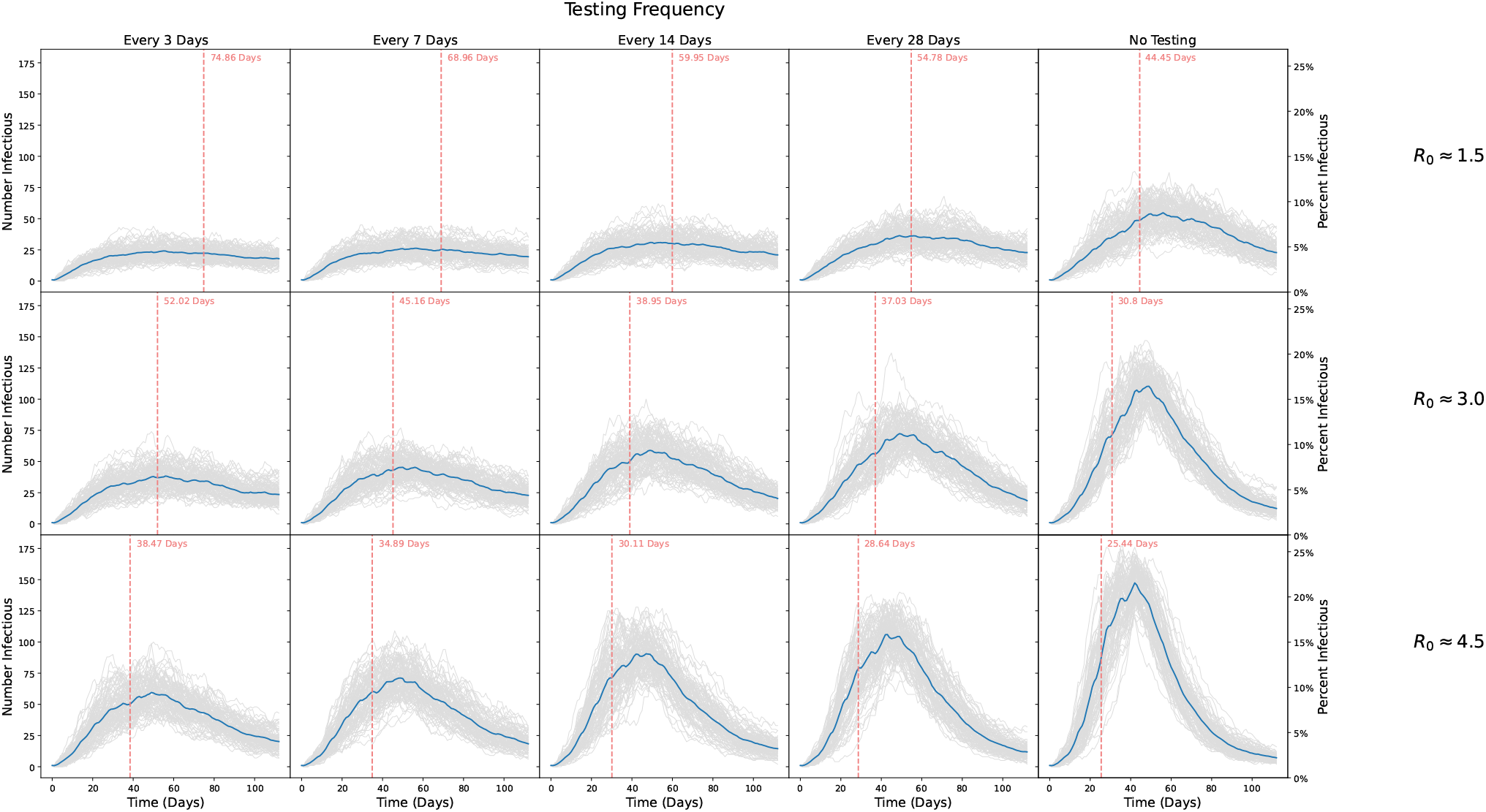
Number of students infected over the course of a simulated 16 week semester. Rows show different epidemic parameters with *R*_0_ ≈ 1.5, *R*_0_ ≈ 3, and *R*_0_ ≈ 4.5, respectively. Columns show scenarios where scheduled testing was done every 3, 7, 14, and 28 days, respectively, and where no testing was done. Grey lines show individual simulations, while blue lines indicate the point-wise average trajectory over all 100 replicates. Vertical red lines and text indicate the average time to reach 20% of the population infected, computed by identifying the time to 20% infected for each realization and averaging those times.

For *R*_0_ ≈ 1.5, testing every 3 days resulted in 202.0 cumulative infections on average (standard deviation 19.1), while testing every 7, 14, and 28 days gave rise to 220.3 (23.5), 251.7 (27.1), and 281.6 (25.8) cumulative infections, respectively. No testing or isolation resulted in 367.9 cumulative infections on average (standard deviation 25.7). For *R*_0_ ≈ 3.0, testing every 3 days resulted in an average of 291.0 cumulative infections (standard deviation 25.9), while testing every 7, 14, and 28 days gave rise to 328.2 (28.2), 380.2 (26.3), and 425.0 (22.7) cumulative infections, respectively. No testing or isolation resulted in 507.7 cumulative infections on average (standard deviation 14.3). Finally, for *R*_0_ ≈ 4.5, testing every 3 days resulted in 375.1 cumulative infections on average (standard deviation 25.9), while testing every 7, 14, and 28 days gave rise to 417.6 (22.8), 466.0 (17.9), and 502.4 (17.1) cumulative infections, respectively. No testing or isolation resulted in 561.4 cumulative infections on average (standard deviation 11.1).

The number of positive tests and isolations for our low, moderate, and high transmission scenarios are shown in Figures 6 and 7, respectively. As expected, more frequent testing leads to more positive tests and isolations. While the number of positive tests mirrors the number of infections, for each scenario testing captures only a fraction of those infected. Notably, we see much more variability in the number of positive tests, with the highest variability seen for the more frequent testing scenarios and for the highest *R*_0_ values.

**Figure 6:**
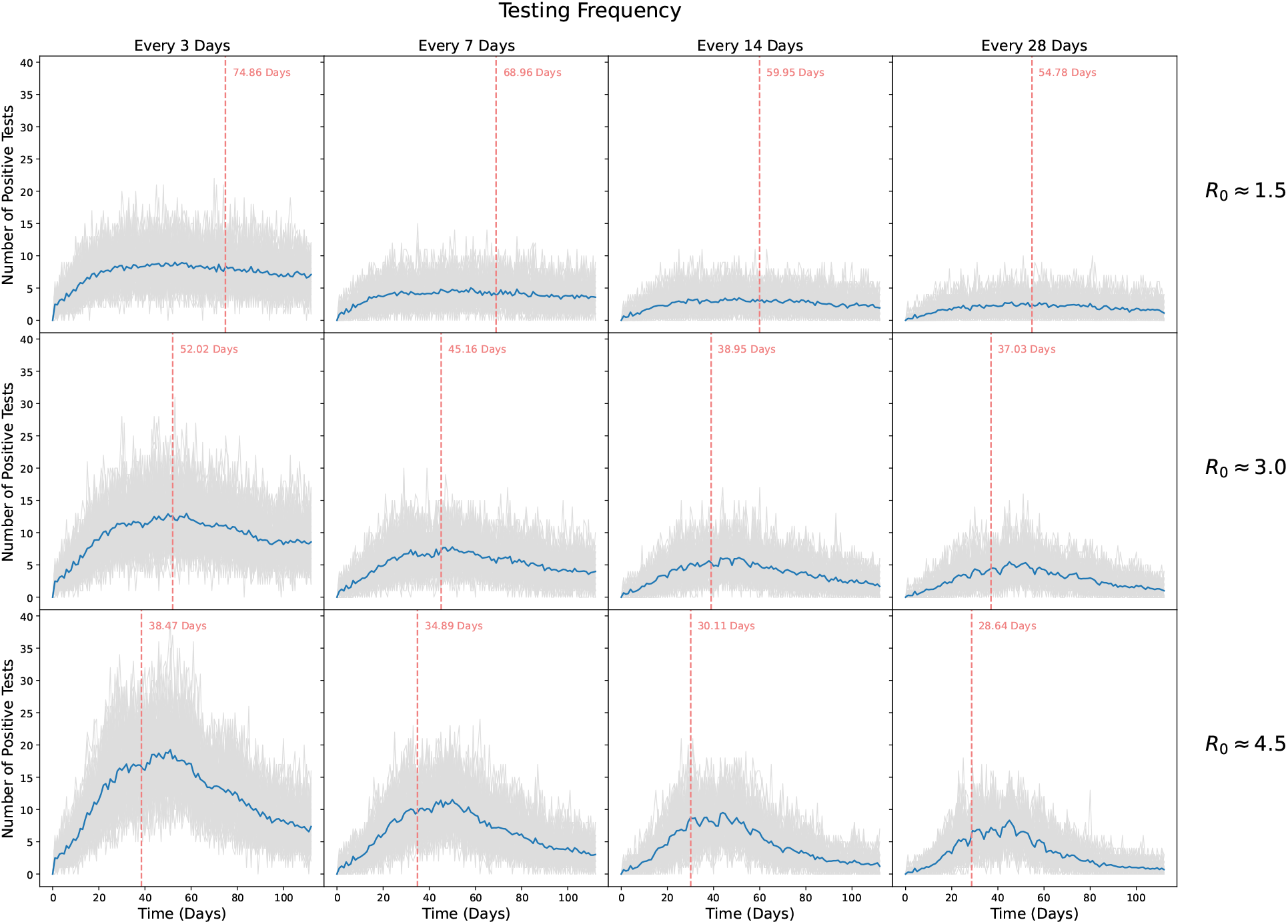
Number of students who tested positive over the course of a simulated 16 week semester. Rows show transmission probabilities of *β* = 0.003078 (*R*_0_ ≈ 1.5), *β* = 0.006157 (*R*_0_ ≈ 3), and *β* = 0.009235 (*R*_0_ ≈ 4.5), respectively. Columns show scenarios where scheduled testing was done every 3, 7, 14, and 28 days, respectively. Grey lines show individual simulations, while blue lines indicate the average trajectory over all 100 replicates. Vertical red lines and text indicate the average time to reach 20% infected.

**Figure 7:**
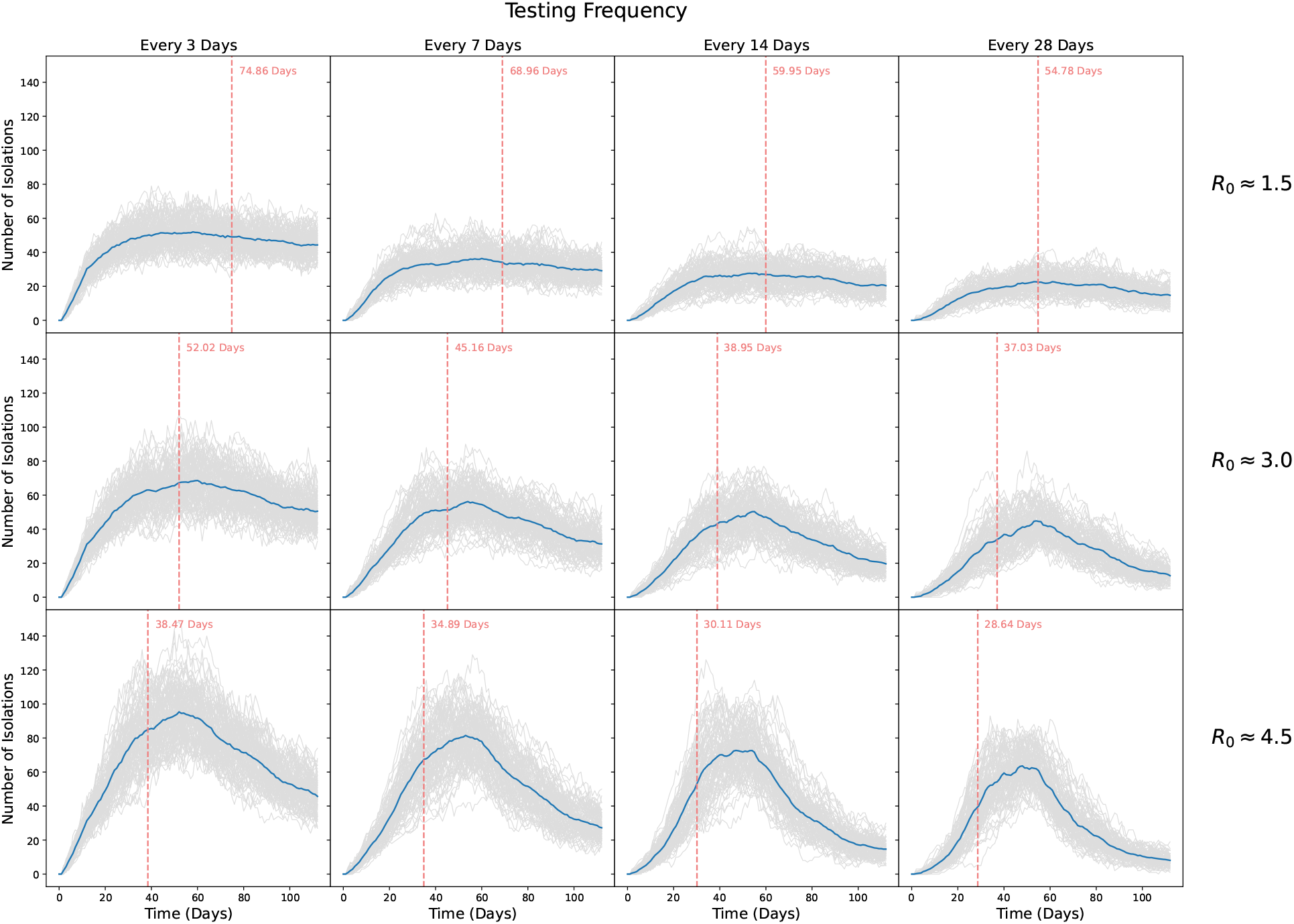
Number of students isolated over the course of a simulated 16 week semester. Rows show transmission probabilities of *β* = 0.003078 (*R*_0_ ≈ 1.5), *β* = 0.006157 (*R*_0_ ≈ 3), and *β* = 0.009235 (*R*_0_ ≈ 4.5), respectively. Columns show scenarios where scheduled testing was done every 3, 7, 14, and 28 days, respectively. Grey lines show individual simulations, while blue lines indicate the average trajectory over all 100 replicates. Vertical red lines and text indicate the average time to reach 20% infected.

As shown in Figure 5, in the low transmission setting the time to infect 20% of all students was 44 days on average without any testing or isolation; testing just once every four weeks increased this time to 55 days, while testing twice a week resulted in 20% of students being infected at day 75 on average. For the moderate transmission setting, it took only 31 days to infect 20% of the student population without testing; testing every four weeks only bought the university 6 additional days, while testing every 3 days led to 20% of students infected at day 52 on average. Finally, under the high transmission scenario, no testing led to 20% of the population infected in less than four weeks, on average at day 25; testing every 28 days only prolonged this by 3 additional days while testing every 3 days bought the university nearly two weeks, but still led to 20% infected by day 38 on average. Hence, testing frequency delayed spread most under the moderate transmission setting, where transmission was high enough for testing to have an impact, but not so high that its efficacy was significantly impeded by delays and compliance issues.

Results for our mask wearing and social distancing scenarios are shown in Figure 8, summarized by relative cumulative incidence. Each cell displays the cumulative incidence with testing divided by the cumulative incidence without testing under comparable levels of mask wearing and social distancing. A value of 1 indicates that testing did not increase or decrease cumulative incidence while a value of 0.5 indicates that testing offered a 50% reduction in cumulative incidence relative to no testing. Testing was most effective at reducing cumulative incidence under low to moderate levels of mask wearing and social distancing since there was more room for improvement. If mask wearing and social distancing were ubiquitous, testing offered only small declines in cumulative incidence since the virus was already well controlled. For a given prevalence of mask wearing and social distancing, testing less frequently offered fewer gains than testing more frequently. Somewhat surprisingly, there was little to no decline in efficacy when mask wearing was clustered on the contact network, perhaps due to the highly connected nature of the network.

**Figure 8:**
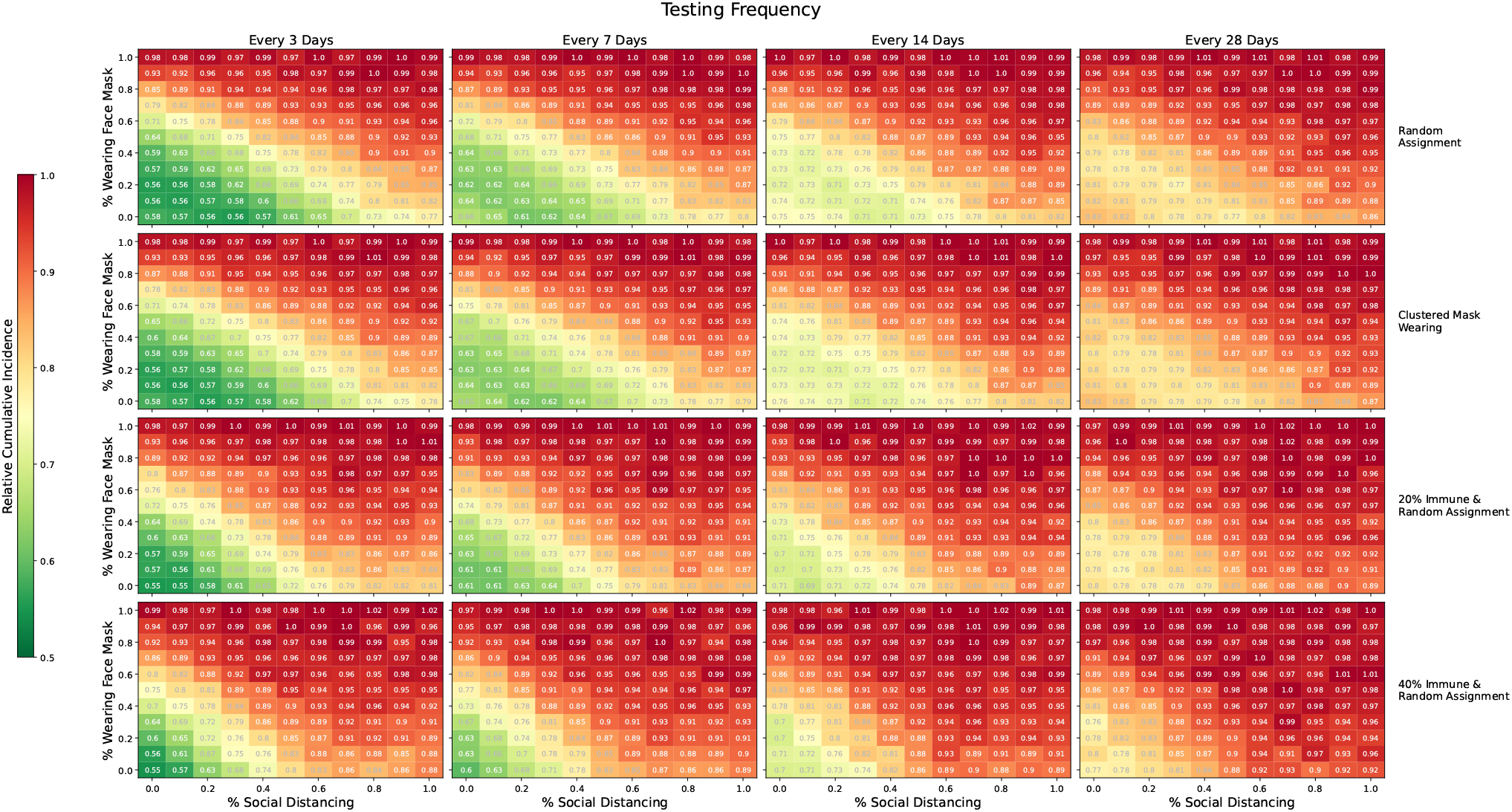
Relative cumulative incidence for various proportions of the population social distancing and/or wearing masks. Each cell displays the cumulative incidence with testing divided by the cumulative incidence without resting under comparable levels of social distancing and mask wearing. Rows one and two show the setting where precautions were randomly assigned to participants and where mask wearing was clustered on the network, respectively. Rows three and four show random assignment of mask wearing and social distancing, but with 20% and 40% of the population immune to SARS-CoV-2 at the outset, respectively. Columns show scenarios where testing was done every 3, 7, 14, and 28 days. In each panel, we consider the moderate transmission scenario with *R*_0_ ≈ 3. A value of 1 indicates that testing offered no reduction in cumulative incidence while a value of 0.5 indicates that testing reduced the cumulative incidence by 50% relative to no testing. All comparisons are for comparable levels of mask wearing and social distancing.

Results from our sensitivity analysis are shown in Figure 9. Under this scenario, with only a 23% reduction in transmission, social distancing had little to no effect on infections. However, despite reduced efficacy, high levels of masking led to sizeable reductions in cumulative incidence. Compared to our original analysis, frequent testing had a much greater impact when masking and distancing were less efficacious. In general, if less than 70% of the population wore masks, frequent testing led to a considerable reduction in infections, regardless of distancing, with more than frequent testing leading to greater gains. Lastly, we considered the setting where a proportion of the population was immune, either due to previous illness or vaccination, results of which are shown in Figure 8. Initial immunity, even in a small fraction of the population, reduced the impact of regular testing. Under no immunity, testing every 3 days offered sizeable benefits if mask wearing and social distancing were below 60%. However, with 20% and 40% immune, testing every 3 days only offered sizeable benefits if mask wearing and social distancing were below 40% and 30%, respectively. Thus, if universities can achieve high levels of mask wearing, social distancing, and/or immunity, our simulations demonstrate they may be able to test less frequently with little or no change in cumulative incidence.

**Figure 9:**
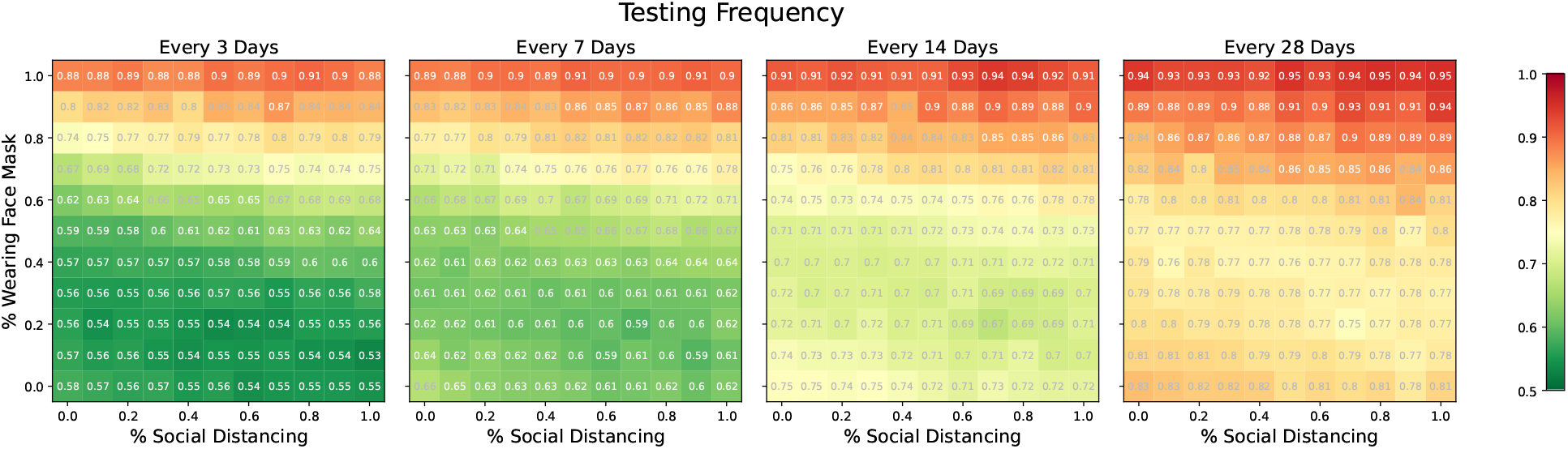
Relative cumulative incidence for various proportions of the population social distancing and/or wearing face coverings with reduced efficacy of masking and distancing. Values are relative to a scenario with comparable proportions of social distancing and mask wearing, but with no testing. Columns show scenarios where testing was done every 3, 7, 14, and 28 days, respectively, with *R*_0_ ≈ 3.

To investigate the comparative effectiveness of testing and isolation, mask wearing, social distancing, and immunity, we conducted a regression analysis of cumulative incidence over all of our moderate transmission simulations (*R*_0_ ≈ 3.0). Cumulative incidence drops by 4.00 or 0.58 per 100 students for every one week increase in testing frequency. Every 10% increase in the proportion of the population social distancing reduces cumulative incidence by 4.56 or 0.66 per 100 students. Likewise, every 10% increase in the proportion wearing masks reduces cumulative incidence by 5.74 or 0.83 per 100 students. Lastly, every 10% increase in the proportion immune decreases cumulative incidence by 17.08 or 2.47 per 100 students. Thus, though testing might be the easiest intervention for colleges to enforce, it also offers the smallest reduction in cases.

## Discussion

When universities abruptly shifted to virtual instruction in the spring of 2020, few anticipated that schools would be grappling with these same challenges nearly a year later. While the past year has given us the promise of effective vaccines and more insight into the dynamics of SARS-CoV-2, schools are still facing an uncertain future. With bumpy vaccine roll-outs and the financial strains of decreased enrollment, many colleges wonder when they will be able to return to some sort of normalcy and what steps they can take to get there sooner ^26^. In this paper, we have endeavored to attenuate some of that uncertainty by examining the efficacy of regularly scheduled (i.e., screening) testing in a residential college population. We found that while testing should be an integral part of every university’s mitigation strategy, steps to reduce transmission among students have a far greater impact. As such, if colleges can achieve low transmission rates, they may be able to relax testing to once or twice a month. While our focus is on a university setting, our methods and results could be applied to other residential environments.

Unlike previous studies, we used a real-world contact network as the basis for our simulation, allowing us to capture the underlying heterogeneous social behavior of college students, which undoubtedly alters how the virus spreads. Additionally, much of the previous work in this area accounted for only a handful of implementation obstacles and compliance issues. Not only do we allow for external infections, a background rate of influenza-like illness, time-dependent test sensitivity, and test result delays, but we also allow for student non-compliance throughout the testing and isolation process.

While our model incorporated many of the challenges we expect to occur on campus, we did not explicitly model contact tracing or vaccine roll-out. We also do not estimate the number of adverse outcomes expected to occur, though deaths and hospitalizations could be approximated from our results if additional assumptions were made. Furthermore, the close proximity data we use dates from well before the pandemic when students were not limiting their interactions with others. Thus, our simulations represent a worst case scenario where students go about their daily lives until they test positive and enter compulsory isolation. However, if universities encourage social distancing, mandate mask-wearing, enhance cleaning protocols, and improve ventilation ^27^, they may be able to reduce interpersonal transmission to the levels we have explored here. These approaches, along with conduct codes, could offer schools a buffer against risky student behaviors.

While this work offers a different look at how repeat testing, isolation, and other strategies can reduce the spread of SARS-CoV-2 on college campuses, it is important to note that there is no one size fits all approach ^28^. The most successful schools will tailor their approach to their specific situation and adapt as circumstances changes. Indeed, the most advantageous strategy may be an agile one where testing frequency is adjusted based on current transmission dynamics, an approach which has yet to be studied rigorously.

## Data and materials availability

Proximity network data from the Copenhagen Network Study are in the public domain (https://doi.org/10.6084/m9.figshare.7267433). All models and code for this project are available through GitHub (https://github.com/onnela-lab/covid-campus).

## Acknowledgments

We thank Marc Lipsitch for his feedback on this project. We also thank Giang T. Nguyen and Max Wang for their critical readings of the manuscript.

## Funding

Harvard University Department of Biostatistics (HH)

U.S. Government scholarship (HH)

MInD: U01 CK000585 (RK)

NIAID R01 award AI138901 (JPO)

## Author Contributions

Conceptualization: JPO

Methodology: HH, RK, JPO

Investigation: HH, RK, JPO

Visualization: HH

Supervision: JPO

Writing – original draft: HH

Writing – review & editing: HH, RK, JPO

